# Assessment of Hydroxychloroquine and Chloroquine Safety Profiles – A Systematic Review and Meta-Analysis

**DOI:** 10.1101/2020.05.02.20088872

**Authors:** Lu Ren, Wilson Xu, James L Overton, Shandong Yu, Nipavan Chiamvimonvat, Phung N. Thai

## Abstract

**Background:** Recently, chloroquine (CQ) and its derivative hydroxychloroquine (HCQ) have emerged as potential antiviral and immunomodulatory options for the treatment of 2019 coronavirus disease (COVID-19). To examine the safety profiles of these medications, we systematically evaluated the adverse events (AEs) of these medications from published randomized controlled trials (RCTs).

**Methods:** We systematically searched PubMed, MEDLINE, Cochrane, the Cochrane Central Register of Controlled Trials (CENTRAL), EMBASE, and the ClinicalTrials.gov for all the RCTs comparing CQ or HCQ with placebo or other active agents, published before March 31, 2020. The random-effects or fixed-effects models were used to pool the risk estimates relative ratio (RR) with 95% confidence interval (CI) for the outcomes.

**Results:** The literature search yielded 23 and 17 studies for CQ and HCQ, respectively, that satisfied our inclusion criteria. Of these studies, we performed meta-analysis on the ones that were placebo-controlled, which included 6 studies for CQ and 14 studies for HCQ. We did not limit our analysis to published reports involving viral treatment alone; data also included the usage of either CQ or HCQ for the treatment of other diseases. The trials for the CQ consisted of a total of 2,137 participants (n=1,077 CQ, n=1,060 placebo), while the trials for HCQ involved 1,096 participants (n=558 HCQ and n=538 placebo). The overall mild or total AEs were statistically higher comparing CQ or HCQ to placebo. The AEs were further categorized into four groups and analyses revealed that neurologic, gastrointestinal, dermatologic, and ophthalmic AEs were higher in participants taking CQ compared to placebo. Although this was not evident in HCQ treated groups, further analyses suggested that there were more AEs attributed to other organ system that were not included in the categorized meta-analyses. Additionally, meta-regression analyses revealed that total AEs was affected by dosage for the CQ group.

**Conclusions:** Taken together, we found that participants taking either CQ or HCQ have more AEs than participants taking placebo. Precautionary measures should be taken when using these drugs to treat COVID-19.

## Introduction

The 2019 coronavirus disease (COVID-19) is caused by the novel, highly infectious, severe acute respiratory syndrome coronavirus 2 (SARS-CoV-2). Since its discovery in December of 2019 in Wuhan, it has now caused a global pandemic. As of March 25, 2020, there were 414,179 confirmed cases and 18,440 deaths from the disease, which brings the mortality to approximately 4.5%.^1^ Thus, scientists, pharmaceutical companies, and medical professionals have united their efforts to develop a vaccine for SARS-CoV-2. Although it is estimated that vaccine development will take at least 12-18 months,^2^ two medications—chloroquine (CQ) and hydroxychloroquine (HCQ)—have emerged as strong contenders to treat the novel COVID-19. CQ and HCQ are antimalarial drugs clinically used to treat inflammatory rheumatic diseases such as rheumatoid arthritis and systemic lupus erythematosus.^3^

Emerging evidence has suggested that these drugs are effective in treating SARS-CoV-2 *in vitro*.^4, 5^ Viral replication begins when the virus attaches to the host cell and penetrates the cell. In the case of SARS-CoV-2, it uses its surface unit (S1) of the S protein to attach to the angiotensin-converting enzyme 2 (ACE2) receptor, which facilitates viral entry.^6^ When African green monkey kidney VeroE6 cells were pretreated for an hour with CQ or HCQ prior to four different multiplicities of infection by SARS-CoV-2, both drugs prevented viral entry as well as post-entry stages of SARS-CoV-2 infection.^4^ Inhibition of viral entry may be due to interference of terminal glycosylation of the ACE2 receptor.^5^ Additionally, CQ and HCQ can alkalinize the phagolysosome, which disrupts the pH-dependent steps of viral fusion and uncoating—processes that are absolutely essential for viral replication.^7^

Moreover, both CQ and HCQ have immunomodulatory properties^3^ that may be beneficial in extreme, life-threatening COVID-19 cases. Indeed, there has been a recent surge in COVID-19 patients with severe hyper immune activity, known as the *cytokine storm syndrome*.^8^ In this patient population, immunosuppression is likely to be beneficial, since the over-active immune response is paradoxically causing more harm than benefit to the patients. Therefore, CQ and HCQ have recently became appealing due to their antiviral and anti-inflammatory properties, which may help treat COVID-19, especially under dire circumstances.

Although the promising findings suggest that CQ and HCQ are great candidates, much concern exists regarding their mechanisms, effective dosing regimen, clinical efficacy, and adverse effects (AEs) with respect to COVID-19. Additionally, precautions must be taken to ensure that the drugs are effective in preventing SARS-CoV-2 viral replication, but do not disrupt the body’s own immune response in fighting against the infection.^9^ Indeed, the rise in popularity of these drugs as potential medications to treat COVID-19 and the current, desperate need for better therapeutics have fueled rapid and ongoing research and clinical trials^10^ to further elucidate their antiviral and anti-inflammatory properties, pharmacodynamics, and safety profiles with respect to COVID-19. Currently, the safety profiles of these drugs are not entirely known due to the lack of large clinical trials, as well as sparse randomized controlled trials (RCTs). Moreover, the drugs have a narrow therapeutic range, which presents another challenge when using these drugs.^11, 12^ Despite these shortcomings, combining the existing data on these drugs can provide powerful and valuable insight regarding their safety profiles, which will not only drive future clinical trials, but also help health professionals make informed decisions.

To this end, our objective for this study is to address the safety profiles of these medications by performing a comprehensive systematic review and meta-analysis of the current published data. We thoroughly searched and screened large databases to acquire RCTs for CQ and HCQ. Since there is a paucity of RCTs for CQ and HCQ for antiviral therapy, we analyzed studies that included the use of CQ and HCQ for non-antiviral treatments. In total, 23 and 14 studies for CQ and HCQ were identified, respectively, that satisfied our inclusion criteria, which are described in the tables. Of these studies, we performed meta-analyses on 6 CQ and 14 HCQ reports, that included a placebo control. Here, we report the results of the meta-analyses to determine the effects of CQ or HCQ on AE relative to placebo control.

## Methods

The meta-analysis was conducted in accordance with the Preferred Reporting Items for Systematic Reviews and Meta-Analyses (PRISMA) guidelines.

### Literature Search and Inclusion Criteria

A comprehensive search strategy was designed to retrieve relevant clinical data from published literature. Our objective was to identify all randomized clinical trials (RCTs) that compared the safety profiles of chloroquine or hydroxychloroquine with placebo or other active agents. We searched PubMed, MEDLINE, Cochrane, the Cochrane Central Register of Controlled Trials (CENTRAL), EMBASE, and the ClinicalTrials.gov for all the RCTs comparing CQ or HCQ with placebo or other active agents, published before March 31, 2020. We also searched conferenced proceedings to acquire relevant papers. Medical subject headings (MeSH terms) and keywords such as “randomized controlled trial,” “adverse effects,” “tolerability,” “toxicity,” and “side effects” were used. This review was not restricted to studies conducted in the English language; it includes reports from any countries that compared CQ or HCQ with placebo or other active agents, since there is a wealth of information in RCTs from many different countries.

Due to the lack of large clinical trials and small numbers of RCTs, we decided to include all the RCTs reporting adverse events (AEs) in patients with different disease conditions, such as rheumatoid arthritis, systemic lupus erythematosus, infectious diseases such as HIV infection, and immune diseases such as Primary Sjögren’s Syndrome. We included all RCTs in adult patients that compared CQ or HCQ with other active agents or placebo.

To be included in the analysis, the study had to fulfill the following criteria: (1) randomized trials which could be open-label, single-blind, double-blind, or parallel group studies; (2) use of CQ or HCQ as one of the interventions; (3) studies comparing CQ or HCQ with placebo or other active agents; and (4) studies should report safety and tolerability data for CQ or HCQ.

Studies were excluded from meta-analysis if: (1) they presented data on children only; (2) they lacked placebo group; (3) study did not present safety and tolerability outcomes; (4) full text could not be sourced; (5) CQ or HCQ was used in combination with other drugs.

### Data Collection and Outcome Measures

Bibliographic details and abstracts of all citations retrieved by the literature search were downloaded to Endnotes X9. All studies were screened and evaluated by two independent reviewers (LR, PNT), which were then checked by a third reviewer (SY). Discrepancies were resolved by discussion in group conferences. Completed data were then thoroughly checked by two additional reviewers (WX, JLO). Data including first author, year of publication, trial design, country where studies took place, purpose of treatment, trial duration, dosage regimen, outcomes and AEs were extracted using a standardized form and presented in table format. Safety evaluation included monitoring of AEs and vital signs. Withdrawals due to AEs were reported.

### Study Quality Assessment and Risk of Bias

Risk of bias in the individual studies included for meta-analysis was assessed using the Cochrane risk assessment tool.^13^ The assessment was performed by two independent reviewers (WX, JLO) and further checked by two additional reviewers (LR, PNT). The completed information is provided in **Supplementary Table 1**.

### Statistical Analysis

Comparison of safety and tolerability outcomes was made between interventions by pooling data from studies using a direct meta-analysis technique. All terminology used when analyzing data was in accordance with the Common Terminology of Clinical Adverse Events handbook. Outcomes were summarized as relative risk ratios. Fixed-effects or random-effects model^14^ were used to pool the risk estimates relative ratio (RR) with 95% confidence interval (CI) for the outcomes. If I^2^ ≤50%, studies were considered homogeneous and fixed-effects model of meta-analysis was used. If I^2^ ≥50%, the heterogeneity is high, so a random-effects model was used. Although we did not alter this in our software output, but I^2^<0% may be considered as I^2^=0%. We analyzed results from RCTs that had placebo controls. Random-effects meta-regression models were used to test whether the relative risk was affected by the age, dosage, or trial duration. Comparisons with no events in either group were excluded. I^2^ statistics was included in all the meta-analyses that were performed, which is a percentage of variance attributed to study heterogeneity. Heterogeneity tests were performed. All analyses were performed using STATA 16 (Stata, College Station, TX, USA).

## Results

### Process of identifying eligible clinical trials

To gather the eligible trials for this systematic review and meta-analysis, we searched through large databases (Pubmed, Cochrane, and ClinicalTrials.gov) and identified those that involved either CQ (n=2,761) or HCQ (n=1,679). Of the published reports we identified, we initially screened them through the titles and abstracts to examine if they were relevant to our objective of identifying safety profiles for CQ and HCQ. Therefore, 134 and 26 reports were initially excluded for CQ and HCQ, respectively. Of the remaining ones (n=70 for CQ and n=80 for HCQ), we performed a more thorough review using the inclusion and exclusion criteria described in the methods. In total, 23 CQ and 17 HCQ studies satisfied our requirements. For our meta-analysis, we extracted data from RCT that had placebo-controlled, rather than studies that compared CQ or HCQ with other drugs. Therefore, a total of 6 studies and 14 studies were used for data extraction for CQ and HCQ, respectively.

### Characteristics of trials, patients, and interventions

We collected data for 23 CQ and 17 HCQ studies. **Table 1** describes the characteristics of the trials, patients, and interventions of CQ, while **Table 2** describes the same parameters for HCQ. The trials indicated with asterisks next to the primary author’s last name were the trials used for our meta-analyses. As manifested from the tables, we did not restrict our systematic review to just the United States. Additionally, investigators used CQ as treatment options for breast cancer (1), malaria (13), hepatitis (3), viral infections (5), and lupus erythematosus (1). To conduct our meta-analysis for CQ, we used 5 double-blinded, placebo-controlled, randomized studies that used CQ for the treatment of breast cancer, autoimmune hepatitis, dengue fever, and influenza. Age of participants ranged from 22- to 57 years-old. Dosing regimen ranged from approximately 107 mg/day to 1000 mg/day. Of these studies, general findings reported in the studies noted that CQ did not have a significant effect when compared with placebo. However, of the studies that compared CQ with other medications, the authors noted that CQ was generally more effective.

**Table 1.**
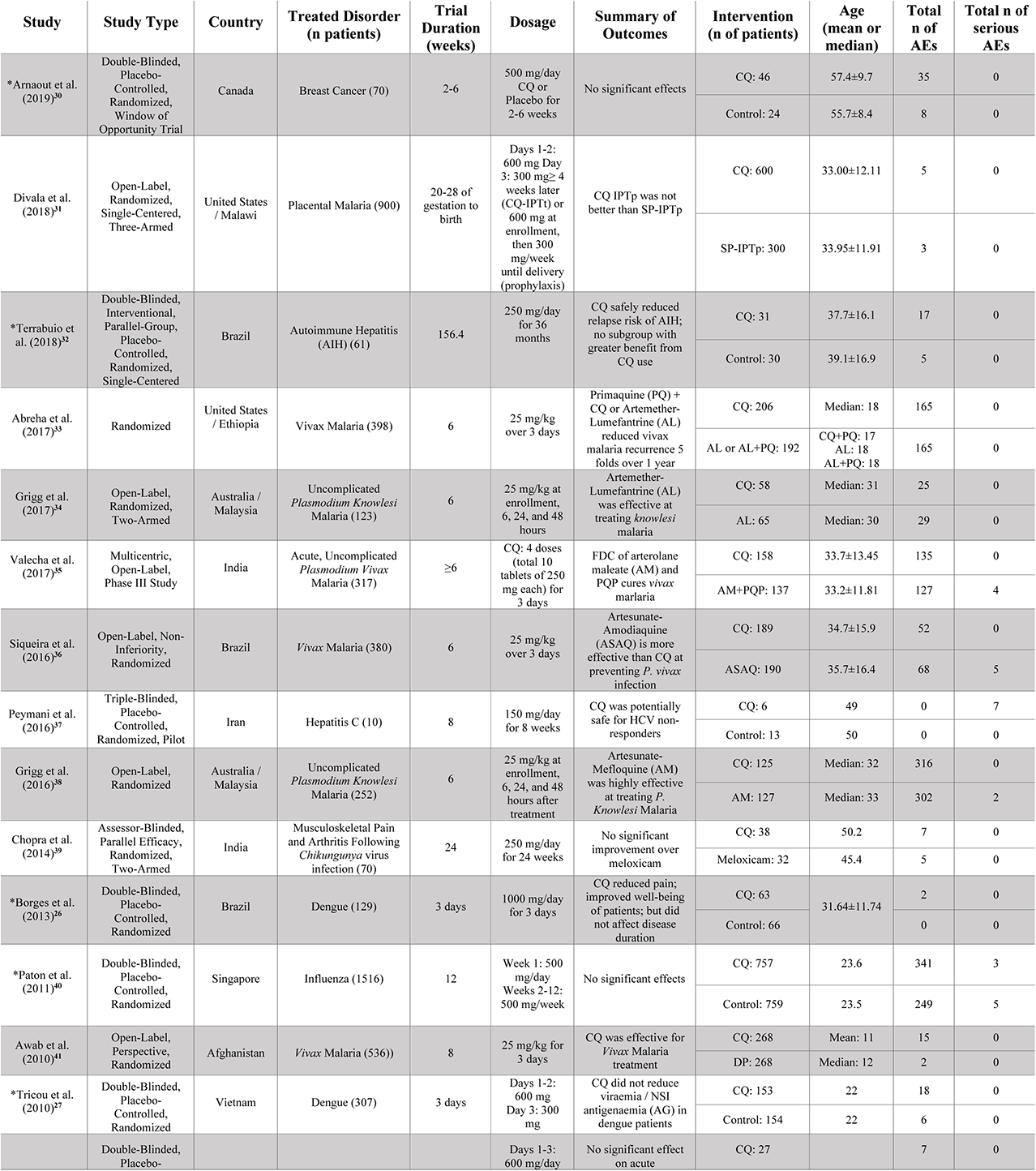
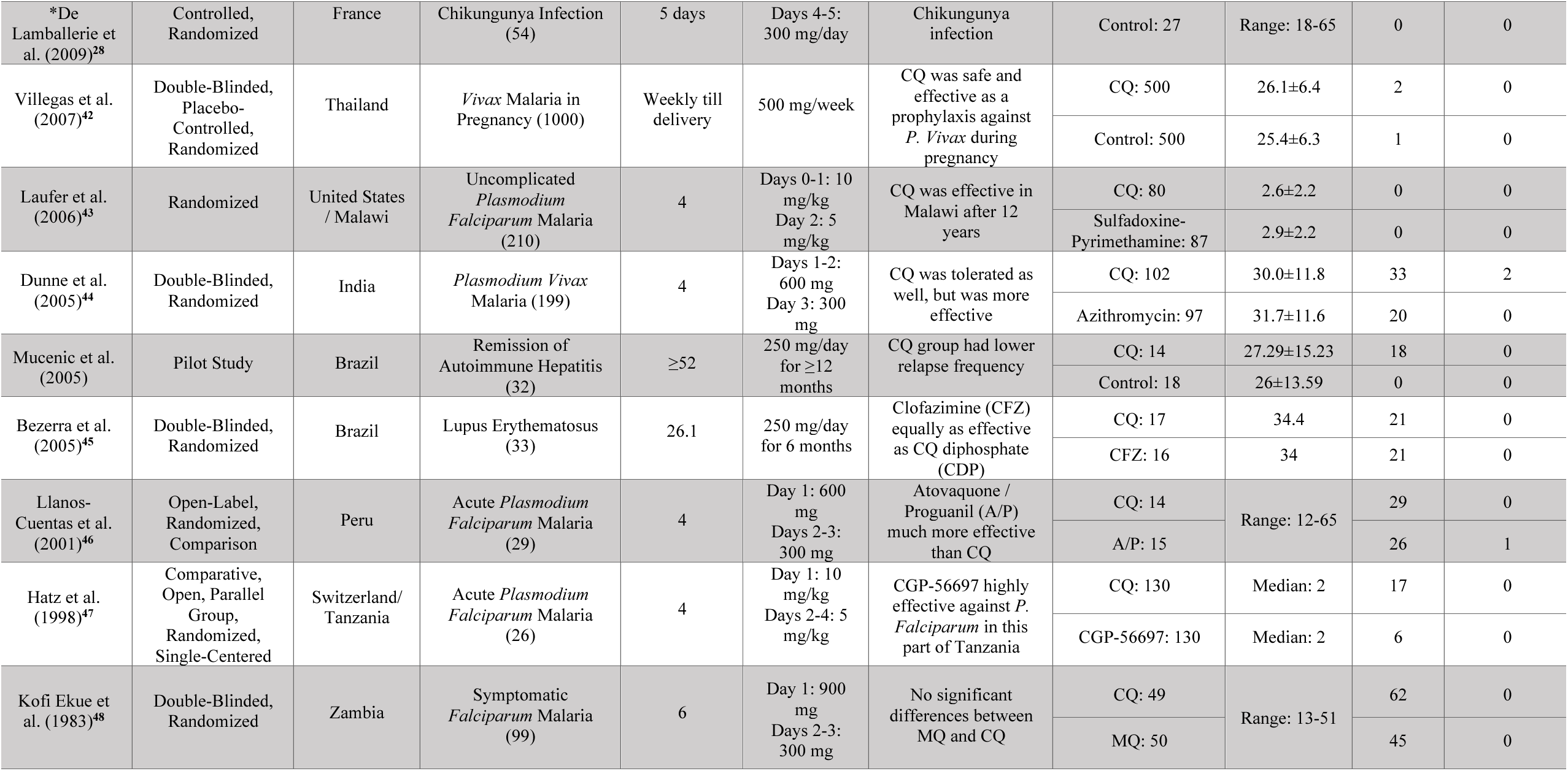
Characteristics of CQ studies

**Table 2.** Characteristics of HCQ studies

| Study | Study Type | Country | Treated Disorder (n patients) | Trial Duration (weeks) | Dosage | Summary of Outcomes | Intervention (n of patients) | Age | Total n of AEs | Total n of serious Aes |
| --- | --- | --- | --- | --- | --- | --- | --- | --- | --- | --- |
| *Chen et al. (2020)<sup>49</sup> | Randomized Pilot Study | China | COVID-19 (30) | 1 | 400 mg/day for 5 days | Prognosis of common COVID-19 patients is good | HCQ: 15 | 50.5±3.8 | 4 | 0 |
|  |  |  |  |  |  |  | Control: 15 | 46.7±3.6 | 3 | 0 |
| *Boonpiyathad et al. (2017)<sup>50</sup> | Single-Blind, Placebo-Controlled, Randomized | Thailand | Anti-Histamine Refractory Chronic Spontaneous Urticaria (CSU) (55) | 12 | 400 mg/day for 12 weeks | HCQ was effective as an adjunct treatment for CSU | HCQ: 46 | 33.00±12.11 | 5 | 0 |
|  |  |  |  |  |  |  | Control: 24 | 33.95±11.91 | 3 | 0 |
| *Wasko et al. (2015)<sup>51</sup> | Double-Blinded, Parallel-Arm, Placebo-Controlled, Randomized | United States | Pre-Diabetes (32) | 13±1 | 400 mg/day for 13±1 weeks | HCQ improved both β-cell function and insulin sensitivity in non-diabetic patients | HCQ: 17 | >18 | 3 | 0 |
|  |  |  |  |  |  |  | Control: 15 |  | 3 | 0 |
| *Gottenberg et al. (2014)<sup>16</sup> | Double-Blinded, Parallel-Group, Placebo-Controlled | France | Primary Sjogren’s Syndrome (120) | 48 | 400 mg/day Placebo or HCQ for 24 weeks, then 400 mg/day HCQ for 24 weeks | No significant effects | HCQ: 56 | 56.3±11.9 | 5 | 5 |
|  |  |  |  |  |  |  | Control: 64 | 55.6±13.9 | 7 | 7 |
| *Solomon et al. (2014)<sup>52</sup> | Blinded, Crossover, Randomized | United States | Rheumatoid Arthritis and Insulin Resistance (30) | 16 | 6.5 mg/kg HCQ or placebo daily for 8 weeks, then crossover to other arm for 8 weeks | No significant change in insulin resistance; minor improvements to total LDL cholesterol | 15 (HCQ → Placebo) | 56±11.4 | 2 | 0 |
|  |  |  |  |  |  |  | 15 (Placebo → HCQ) | 56±11.4 | 0 | 0 |
| *Rotaru et al. (2014)<sup>17</sup> | Randomized, Pilot, Triple Masking | United States | Kidney Failure, Chronic Cardiovascular Disease Arteriosclerosis (8) | 25 | 200 mg/day for 10 days±4 days, then 200 mg twice daily for 6 months | Terminated (Lack of Funding) | HCQ: 7 | 18-65: 4 >65: 3 | 2 | 0 |
|  |  |  |  |  |  |  | Control: 1 | 18-65: 1 | 0 | 0 |
| *Paton et al. (2012)<sup>53</sup> | Double-Blinded, Randomized, Placebo-Controlled | United Kingdom | HIV (83) | 48 | 400 mg/day for 48 weeks | No significant effects | HCQ: 42 | 37.1±7.7 | 41 | 0 |
|  |  |  |  |  |  |  | Control: 41 | 38.3±10.8 | 26 | 0 |
| *Fong et al. (2007)<sup>54</sup> | Double-Blinded, Placebo-Controlled, Randomized | United States | Chronic Graft-Versus-Host Disease (95) | 55 | 121 days at 800 mg/day | No effects | HCQ: 46 | 48 | 1 | 0 |
|  |  |  |  |  |  |  | Control: 49 | 46 | 1 | 0 |
| *Gerstein et al. (2001)<sup>55</sup> | Double-Blinded, Placebo-Controlled, Randomized | Canada | Type 2 Diabetes Mellitus (135) | 78.2 | 300 mg first month, 450 mg second, and 600 mg third, daily | HCQ improved glycemic control in patients with poorly controlled type 2 diabetes | HCQ: 69 | 57.5 | 3 | 0 |
|  |  |  |  |  |  |  | Control: 66 | 57.5 | 1 | 0 |
| *Van Gool et al. (2001)<sup>56</sup> | Double-Blinded, Parallel-Group, Multicenter | The Netherlands | Dementia in Early Alzheimer’s Disease (168) | 78.2 | <65 kg: 200 mg/day >65 kg: 400 mg/day; 18 months | No significant effects | HCQ: 83 | 70.4±8.3 | 20 | 5 |
|  |  |  |  |  |  |  | Control: 85 | 70.7±8.5 | 15 | 2 |
| *Sperber et al. (1995)<sup>57</sup> | Double-Blinded, Placebo-Controlled, Randomized | United States | HIV-1 (40) | 8 | 800 mg/day for 8 weeks | HIV-1 RNA declined significantly in the HCQ group over 8 weeks; increased in placebo group | HCQ: 19 | 39.1±6.6 | 0 | 0 |
|  |  |  |  |  |  |  | Control: 19 | 40.6±12.5 | 0 | 0 |
| *The HERA Study Group (1995)<sup>58</sup> | Double-Blinded, Placebo-Controlled, Randomized | Canada | Early Rheumatoid Arthritis (120) | 36 | 200 mg/day for 2 weeks. If no side effects, 400 mg/day | Improved pain and disability of recent arthritis | HCQ: 59 | 53±13.5 | 25 | 1 |
|  |  |  |  |  |  |  | Control: 60 | 53±14.8 | 19 | 0 |
| *Clark et al. (1993)<sup>59</sup> | Double-Blinded, Placebo-Controlled, Randomized | Mexico | Early Rheumatoid Arthritis (126) | 24 | 400 mg/day for 24 weeks | HCQ effectively improved early rheumatoid arthritis | HCQ: 65 | 39 | 28 | 0 |
|  |  |  |  |  |  |  | Control: 65 | 36 | 28 | 1 |
| *Kruize et al. (1993)<sup>60</sup> | Double-Blinded, Crossover, Placebo-Controlled | The Netherlands | Primary Sjogren’s Syndrome (19) | 52.2 | 400 mg/day for 12 months | No significant effects | 10 (HCQ → Placebo) | 52.8±16.1 | 0 | 1 |
|  |  |  |  |  |  |  | 9 (Placebo → HCQ) | 51±15.8 | 0 | 0 |
| Fries et al. (1993)<sup>61</sup> | Observative | United States | Rheumatoid Arthritis (2747) |  |  | HCQ was the least toxic drug tested |  |  | 0 |  |
|  |  |  |  |  |  |  |  |  | 0 |  |
| Faarvang et al. (1993)<sup>62</sup> | Double-Blinded, Multicenter, Parallel-Group, Placebo-Controlled, Randomized | Denmark | Rheumatoid Arthritis (91) | 26.1 | 250 mg/day HCQ and 2g/day Placebo OR 250 mg/day + S for 6 months | HCQ and Sulphasalazine (S) had no improvement over HCQ alone | 62 (HCQ + Placebo & HCQ + Sulphasalazine) | 61 | 7 | 0 |
|  |  |  |  |  |  |  | 29 (Placebo + Sulphasalazine) | 61 | 0 | 0 |

Similarly, the 17 HCQ studies (**Table 2**) that we examined were conducted from a plethora of countries and used HCQ to treat a myriad of disorders, which included dermatologic disorders (1), rheumatoid arthritis (5), HIV (2), Primary Sjögren’s Syndrome (2), graft-versus host disease (1), diabetes (2), chronic spontaneous urticaria (1), dementia (1), kidney failure (1), cardiovascular disease (1), and COVID-19 (1). To conduct our meta-analysis for HCQ, we used RCTs that were pilot studies (one specifically for COVID-19), 1 single-blinded, and the rest double-blinded. In the table, these studies have asterisks next to the primary author’s last name. For these particular reports, age of participants ranged from 33 to 70 years. Dosage schedule ranged from 200 mg/day to 800 mg/day, with a mode of 400 mg/day. General outcomes from about a third of the studies revealed that HCQ had no significant effects, while the rest of the studies showed that it was effective for the disorders.

### Mild, severe, total AEs, and withdrawals due to AEs from trials involving CQ and HCQ

The CQ meta-analyses of mild, serious, total AEs, and withdrawals due to AEs were based on 6 comparisons between CQ and placebo (control), while the HCQ meta-analyses of mild, serious, total AEs, and withdrawals due to AEs were based on 14 comparisons between HCQ and placebo (control), as depicted in **Figure 2**. When assessing mild AE (**Figure 2A**), the overall relative risk (RR) of CQ compared with placebo was 2.15 (95% CI 1.37-3.37, p<0.01), while the overall RR of HCQ compared with placebo was 1.27 (95% CI 1.05- 1.55). The RR for severe AEs (**Figure 2B**), however, was insignificant for both drug usage when compared with placebo. When assessing total AEs of either drug compared with placebo (**Figure 2C**), the combined RR for CQ was 2.25 (95% CI 1.40-3.62, p<0.01), while for HCQ it was 1.28 (95% CI 1.05-1.54, p<0.05). There was statistical evidence of overall heterogeneity between CQ trials with regards to total AEs (I^2^ = 55.79%). Withdrawals due to AEs was only significant with CQ compared with placebo. As evident in **Figure 2D**, the overall RR was 2.33 (95% CI 1.17-4.64, p<0.05). There was no evidence of heterogeneity (I^2^ = 0%). Taken together, these data suggest that both drugs induced higher mild and total AEs as compared to control. Additionally, CQ had significantly more patients that withdrew from the studies due to AEs.

**Figure 1.**
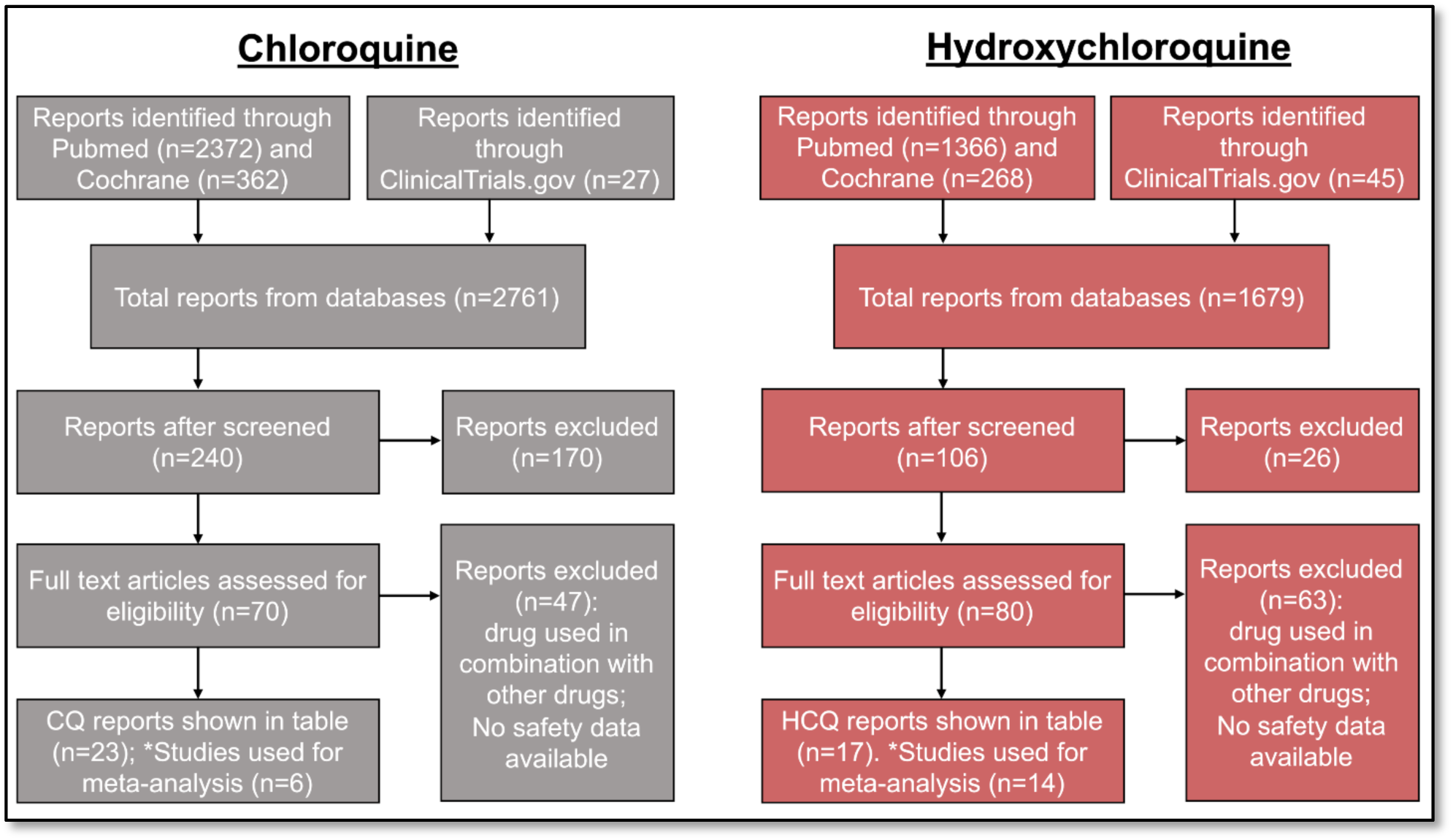
Process of identifying eligible clinical trials for CQ and HCQ. Reports were identified through Pubmed, Cochrane, and ClinicalTrials.gov. We used the same process of study collection for both CQ and HCQ. We performed an initial screening, followed by a more stringent screening using our selection criteria. The studies that remained after all the exclusion were the ones used for data extraction. In total, we identified 23 and 17 studies for CQ and HCQ, respectively, which are described in Tables 1 and 2. Of those studies, 6 CQ and 14 HCQ reports are placebo-controlled RCTs, so we used these studies for our data analysis.

**Figure 2.**
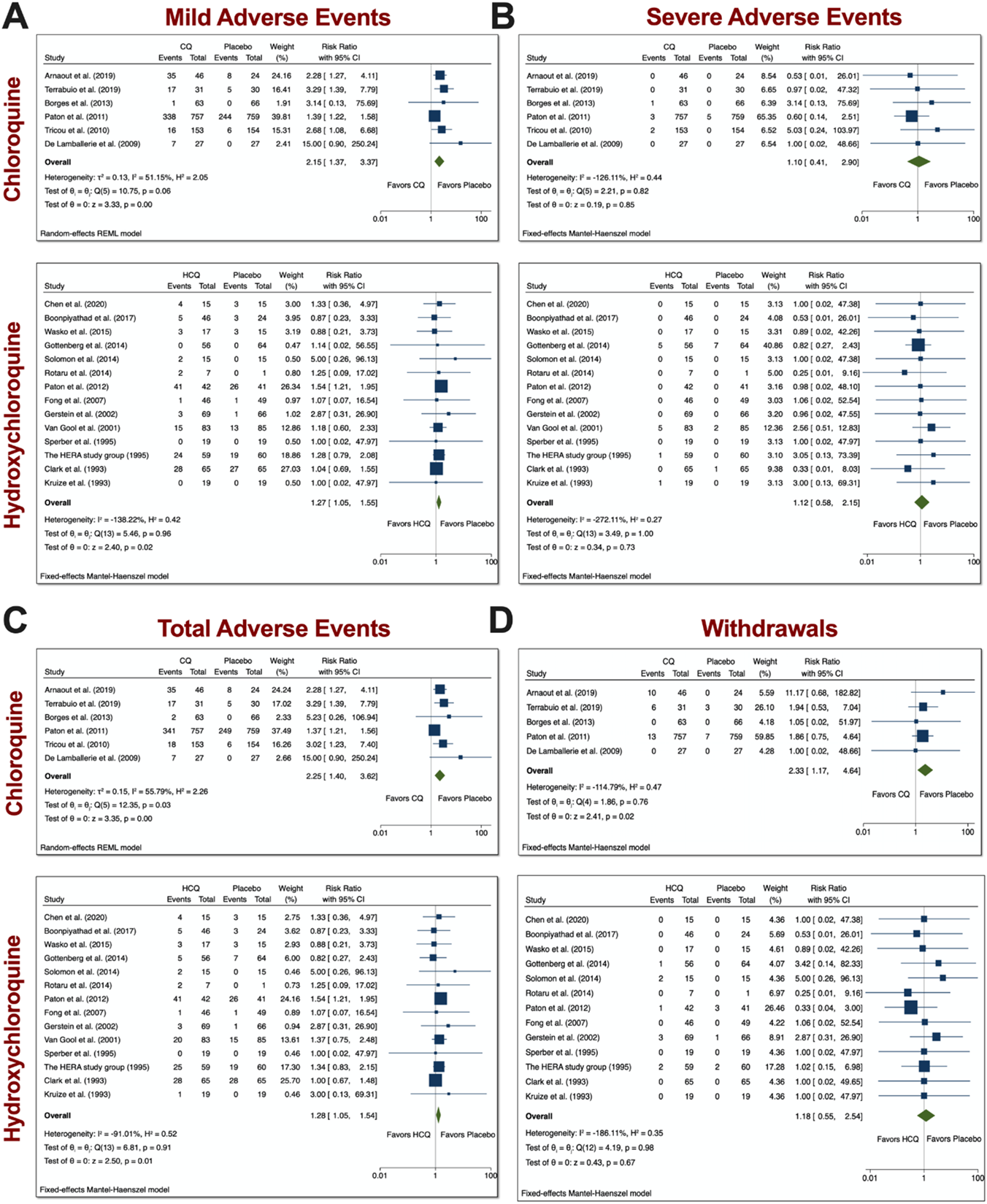
Mild, severe, total AEs, and withdrawals due to AE from trials involving CQ and HCQ. We performed 6 comparisons between CQ and placebo and 14 comparisons between HCQ, as evident in the forest plots. AEs were divided into **A**) mild, **B**) severe, and **C**) total. **D**) Additionally, we also examined withdrawals from trials due to AEs. Meta-analyses were performed. We tested heterogeneity between trials, as well as overall effect. Statistical data are displayed in the forest plots.

### System analyses from trials with CQ and HCQ

Based on the reported AEs, we divided our analyses to examine four groups: neurologic, gastrointestinal (GI), dermatologic, and ophthalmic AEs. Neurologic AEs reported by participants included headache, dizziness, neuropathy/seizure, or other central nervous system (CNS) related AEs; GI AEs included vomiting, nausea, abdominal pain, diarrhea, liver dysfunction, or non-specific GI AEs; dermatologic AEs included rash, itchiness, dryness; and ophthalmic AEs included blurred vision or pain. With the usage of CQ, there was a significant increase in all four groups of AEs (**Figure 3**). The overall RR was 2.75 (95% CI 2.14-3.52, p<0.01) for neurologic AEs; 2.86 (95% CI 2.08-3.95, p<0.01) for GI AEs; 1.92 (95% CI 1.14, 3.26, p<0.05) for dermatologic AEs; and 5.76 (95% CI 2.25-14.74) for ophthalmic AEs. No heterogeneity between the trials were observed. With the usage of HCQ, there was no significance increase in the groups that we examined. These data suggest that patients treated with CQ experienced more neurologic, dermatologic, ophthalmic, and GI AEs relative to placebo control.

**Figure 3.**
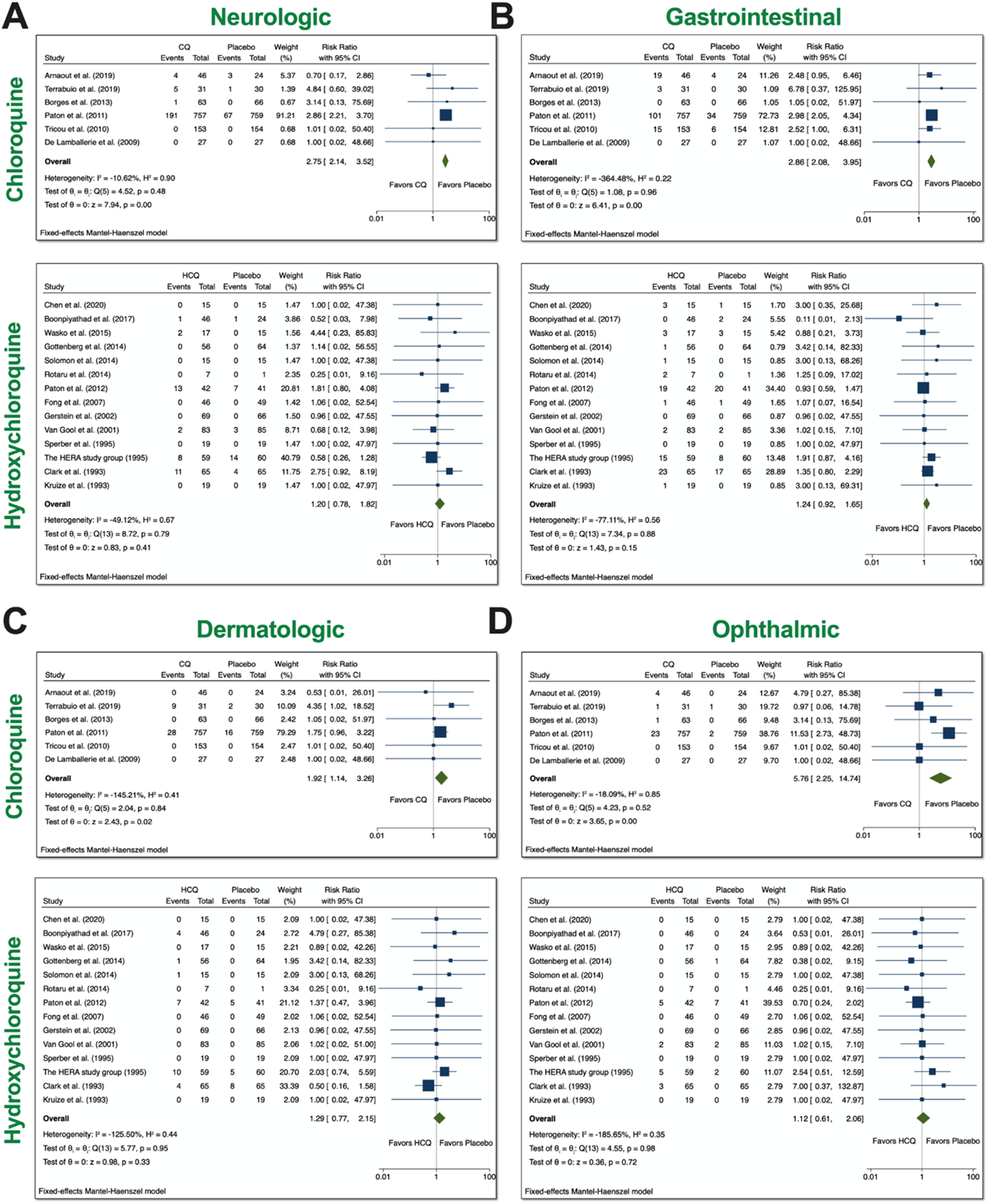
System analyses from trials with CQ and HCQ. We performed 6 comparisons between CQ and placebo and 14 comparisons between HCQ, as evident in the forest plots. AEs were divided into 4 groups: **A**) neurologic, **B**) gastrointestinal (GI), **C**) dermatologic, **D**) and ophthalmic AEs. Using meta-analyses, we tested heterogeneity between trials, as well as overall effect. Statistical data are displayed in the forest plots.

Further analyses on heterogeneity, as well as publication bias, can be seen in **Supplementary Figures 2-5**.

### Stratification of all AEs

To fully appreciate the wealth of information from the RCTs, we constructed a flow chart that contains information on the number of participants who experienced a certain AE, as well as the percentages. Four groups (CNS, GI, skin, and vision) underwent meta-analyses (**Figure 3**), since they had robust reports in the studies that we examined. In **Figure 4**, panel A and B show the charts for CQ and HCQ, respectively. The 6 CQ studies contained a total of 1,077 participants for CQ-treated group and it contained a total of 1,060 participants for placebo-treated. Of these participants, 435 (40.4%) and 270 (25.5%) AE were reported in the CQ and placebo group, respectively. The highest reported AEs for the CQ group occurred in the CNS, with about 18.7% of overall CQ participants reporting headache, dizziness, neuropathy, or other CNS-related AEs. In contrast, placebo group had higher reports for respiratory distress, such as coughing, sore throat, or running nose.

**Figure 4.**
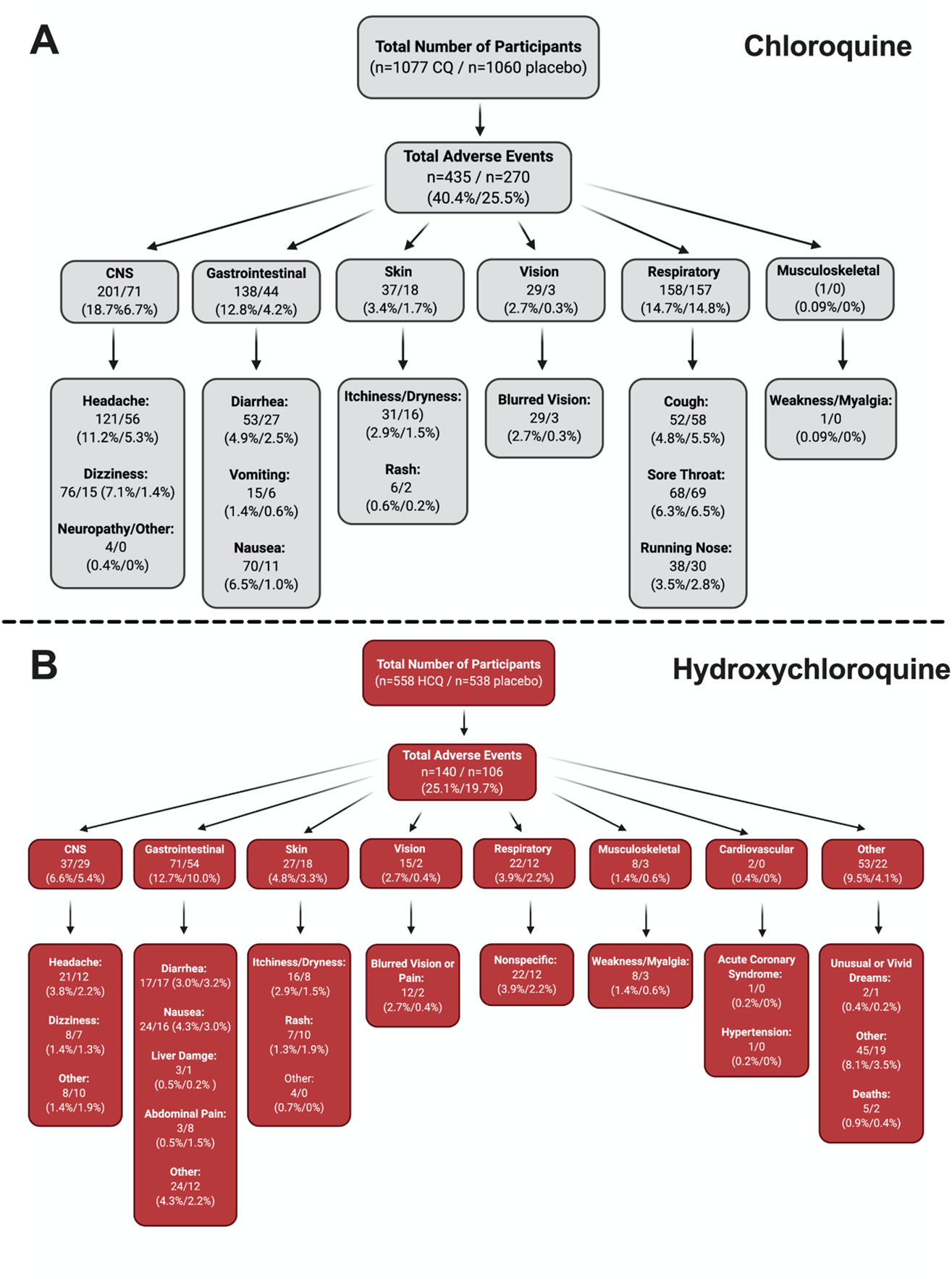
Stratification of all AE. To fully appreciate the wealth of data regarding CQ and HCQ AE, we divided the AE into different categories. Panel **A**) depicts the data for CQ, while panel **B**) shows the data for HCQ. Both panels begin with the total number of participants in the studies (n=6 CQ, n=14 HCQ), which is then followed by the total number of AE. The AE were then divided into different systems, which is then broken down into specific AE. Figure was generated using BioRender.

The 14 HCQ studies contained 558 participants for HCQ-treated group and 538 participants for placebo-treated group. Total AEs reported for HCQ was 140 (25.1%), while total AEs reported for placebo was 106 (19.7%). Gastrointestinal AEs, such as diarrhea, nausea, liver damage, abdominal pain, and other non-specific GI AEs seemed to be the most dominant for both groups. Interestingly, cardiovascular AEs was reported in 2 of the studies that we examined; one AE involved hypertension, while the other was acute coronary syndrome. Together, these stratified data provide ample information regarding the percentage of participants who experienced specific AEs.

### Subgroup meta-analysis for CQ and HCQ with respect to age, duration, and dosage

Since we found a significant increase in total AEs when taking either drugs, we tested whether differences in age, duration, or dosage had any bearing on the results. We therefore performed subgroup meta-analysis. First, we examined age (**Figure 5A**). We divided the CQ trials into two groups: participants <30 years-old and participants ≥30 years-old. We stratified the HCQ trials into two groups: participants <50 years-old and participants ≥50 years-old. These ages were chosen to ensure that there was robust comparison, since the number of RCTs was very limited. We found that there was no group difference in either case, which suggests that age (younger vs older) had no bearing on the total AEs experienced in participants.

**Figure 5.**
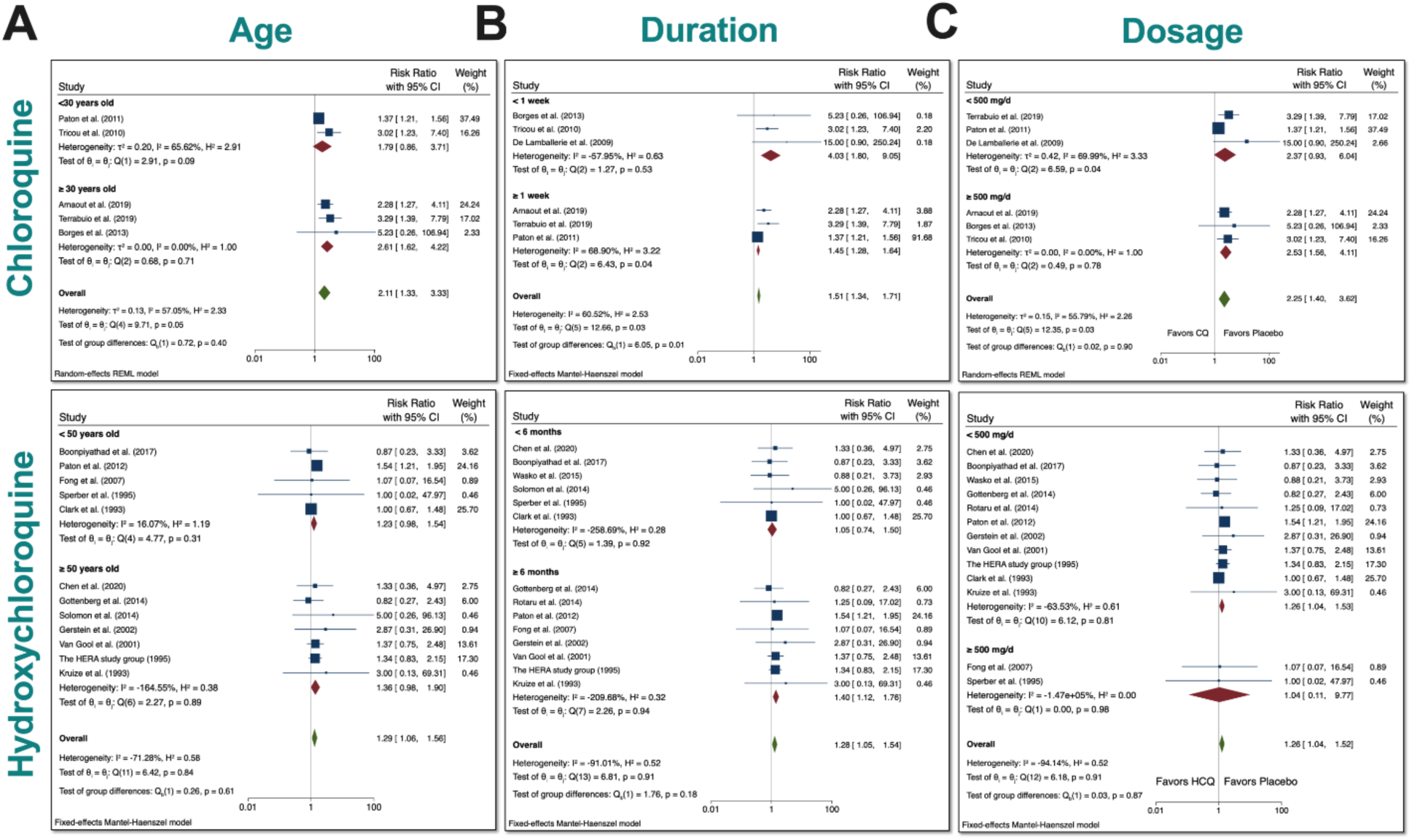
Subgroup meta-analyses for CQ and HCQ with respect to age, duration, and dosage. We stratified the dosages used in the studies for both CQ and HCQ into two subgroups. We then performed subgroup analysis for dosage and trial duration. **A**) For age, we separated CQ trials into <30 years-old and ≥30 years-old, while we separated HCQ trials into <50 years-old and ≥50 years-old. **B**) For drug duration, we divided CQ studies into <1 week and ≥1 week, while we divided HCQ studies into <6 months and ≥6 months. **C**) And for dosage, we wanted to investigate if there was a difference in using <500 mg/day versus using ≥500 mg/day for either drug. Statistical data are presented in the figures.

Next, we assessed whether duration had any relevance to total AEs (**Figure 5B)**. CQ trials were divided into two groups: <1 week and ≥1 week. HCQ trials were divided into two groups: <6 months and ≥6 months. There was significance difference between the two groups for CQ (p<0.05). The overall RR of total AEs in trials that lasted <1 week was 4.03 (CI 95% 1.80-9.05), while the overall RR of total AEs in trials that lasted ≥1 week was 1.51 (CI 95% 1.34-1.71). Additionally, there was evidence of heterogeneity (I^2^=60.52%) between the two groups. Although overall heterogeneity did exist between the groups, it is important to note that when these studies were separately analyzed, there was statistical significance for either group (p<0.05). Upon close inspection of HCQ trials, we noted that they generally lasted longer. There was no evidence that a higher duration is any different than a lower duration in terms of RR of total AEs.

Finally, to determine if there were significant differences between a low versus a high dosage with respect to total AEs for either drug, we stratified the dosages into two groups: <500 mg/day and ≥500 mg/day. This arbitrary grouping ensured that we included enough studies in each group for CQ, since the number of RCT for CQ is limited. Additionally, most of the HCQ studies used 400 mg/day as the dosage (8 HCQ studies), so many reports fell below the <500 mg/day threshold that we set. As evident in our meta-analyses, there was no significant difference in the subgroups when using either dosage for both drugs.

Taken together, there was no statistical evidence to suggest that (younger vs older) age and (lower vs higher) dosage differentially affected the total AEs when using either drug. Additionally, there was statistical evidence to suggest that separation according to duration had an impact on total AEs in the CQ studies.

### Meta-regression analyses for CQ and HCQ

Meta-regression analyses were performed to determine the relationship between RR and age, duration of trial, and dosage, as depicted in **Figure 5**. We examined if age of participants, duration of trial, or dosage has any effects on total AEs or withdrawals due to AEs. The size of the symbols indicates more weight towards a particular study. In all plots, the predicted regression lines and 95% confidence-interval lines are displayed. Regression of logarithm of RR of total AE with CQ and dosage revealed that dosage had an effect on total AEs. Age and duration of trial did not affect the total AEs for CQ. For HCQ, there was no evidence that age, duration of trial, or dosage affected total AEs. Further meta-regression analyses can be found in **Supplementary Figure 1**.

## Discussion

The current pandemic with SARS-CoV-2 has relentlessly claimed thousands of lives and caused significant economic hardship. The current need for viable therapeutic options while vaccine development is in progress has resulted in the proposal of numerous antiviral medications.^15^ Chloroquine (CQ) and its derivative hydroxychloroquine (HCQ) have been circulating in the media as potential drugs to treat COVID-19. However, little is known regarding their safety profiles due to the lack of randomized controlled trials (RCTs). To address this urgent issue, we performed a systematic review and meta-analysis by pooling the existing published data of adverse events (AEs) for CQ and HCQ relative to placebo.

After comprehensively perusing through the literature, we identified 23 CQ and 17 HCQ eligible studies that satisfied our inclusion criteria (**Figure 1**). The characteristics of these studies are documented in **Tables 1 and 2**. For our meta-analyses, we included 6 CQ and 14 HCQ, since these studies were placebo-controlled. We found that the usage of either drug increased the relative risk (RR) for mild and total AEs (**Figure 2**). Further system analyses showed that overall participants in the CQ trials experienced more neurologic, gastrointestinal, dermatologic, and ophthalmic AEs (**Figure 3**). We did not observe a significant increase in the HCQ group compared to placebo with regards to these AE categories. Therefore, the significant increase in mild and total AEs from HCQ was most likely attributed to other reported AEs, such as respiratory and cardiovascular AEs.

Given the severity of cardiovascular AEs, it is critical to note that two studies reported two cardiovascular AEs: one was hypertension and the other was acute coronary syndrome.^16, 17^ Although there were no cardiovascular AEs reported in the CQ studies that we analyzed, its cardiotoxicity has also been noted in a plethora of studies.^18^ An excellent systematic review article by Chatre et al. documented the cardiac complications that are attributed to CQ and HCQ.^18^ In their review, they found that among other cardiovascular complications, conduction bundle or atrioventricular block were reported more frequently. Moreover, QT interval prolongation has been noted in numerous studies.^19–22^ Severely prolonged QT interval can lead to lethal arrhythmias and sudden cardiac death. Therefore, the prevalence of these cardiovascular AEs warrants periodic electrocardiogram (ECG) monitoring when participants are undergoing these therapies, as cardiovascular AEs can be fatal. Since many of the studies that involved participants who experienced cardiovascular complications were not RCTs with placebo controls, we were unable to include them in our meta-analyses.

Overall, participants who took CQ exhibited more AEs (40.4%) relative to placebo control (25.5%, **Figure 4**). In the HCQ studies, only 25.1% of total AEs were reported versus 19.7% for placebo. The high percentage of total AEs occurring with CQ participants is concerning, but consistent with the consensus that HCQ is a safer alternative to CQ.^4, 23–25^ When total AEs were stratified according to different organ systems, we found that CQ had more participants exhibiting CNS AEs (18.7%), while HCQ participants had more participants experiencing GI AEs (12.7%). It is worth noting that only 6.6% of HCQ participants exhibited CNS AEs. The extra hydroxyl group in HCQ may decrease the occurrence of CNS AEs. More mechanistic, controlled studies need to be performed to confirm this finding.

Furthermore, subgroup analyses (**Figure 5**) revealed no evidence in differences of RR of total AEs when studies were divided by age (younger vs older) and dosage (lower vs higher). However, when we performed meta-regression analyses (**Figure 6**), there was a relationship between dosage and total AEs in the CQ group, which suggests that the subgroup meta-analyses for dosage would be more robust if more CQ RCTs existed.

**Figure 6.**
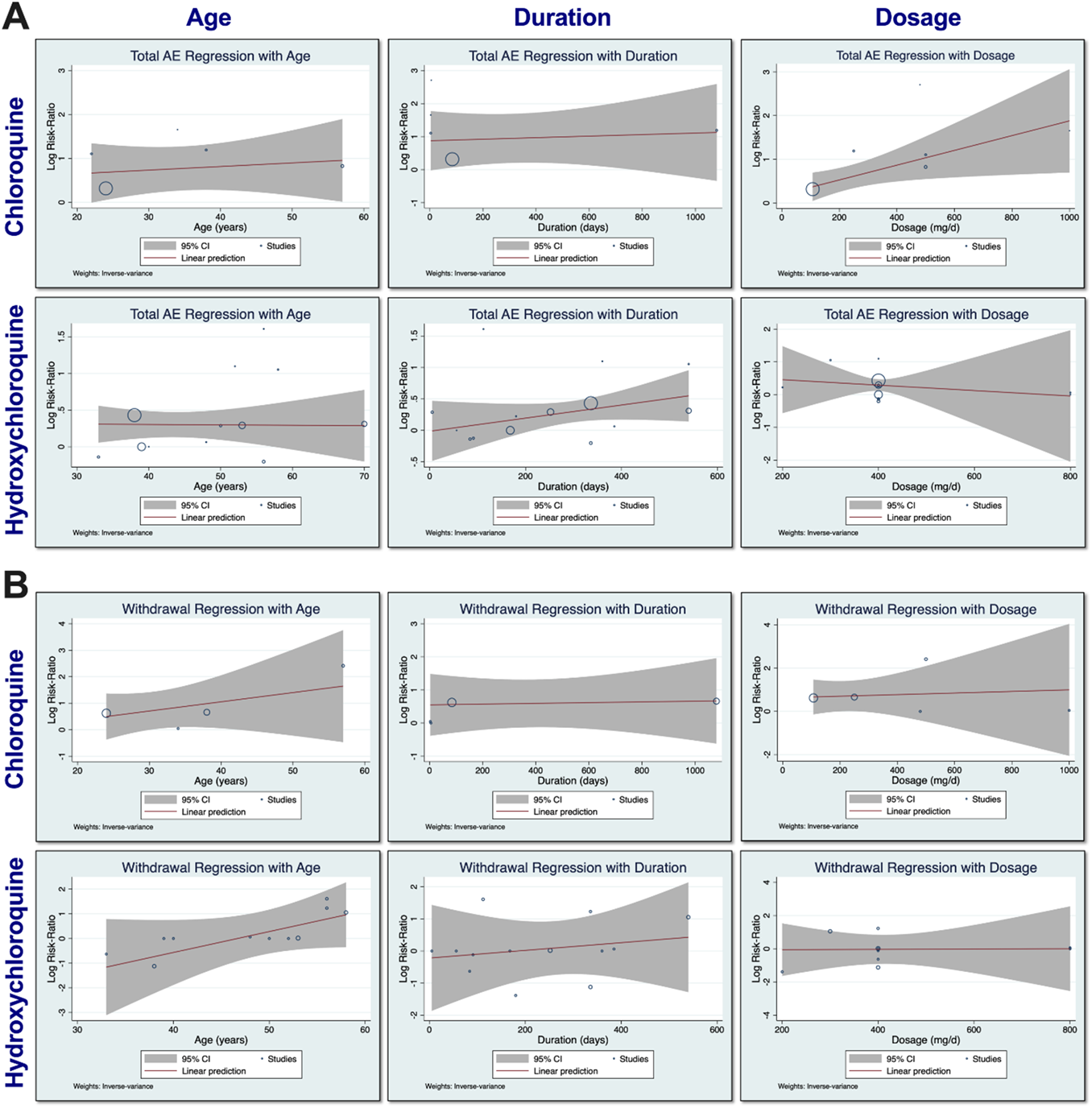
Meta-regression analyses for CQ and HCQ. Meta-regression analyses were performed using the log RR of total AEs with respect to age, duration, and dosage for both CQ and HCQ, as shown in panel **A**). Panel **B** depicts the meta-regression analyses of RR of withdrawals due to AEs with respect to age, duration, and dosage for both CQ and HCQ. Statistical data are presented in the figure.

Interestingly, we found that duration of drug had an impact on total AEs when using CQ, with lower duration (<1 week) having higher RR. When examining studies with duration <1 week,^26–28^ we noticed that CQ was used to treat Dengue (2) and Chikungunya (1) infections. CQ was used to treat various other conditions in trials with higher duration (≥1 week). Upon closer inspection of the HCQ duration subgroup analysis, we observed that studies with duration <6 months had an insignificant overall RR of total AEs, while studies with duration ≥6 months had a very significant overall RR of total AEs (p<0.01). However, when these two groups were compared (p=0.18), there was no significant difference between them. Indeed, given the long half-life of HCQ,^29^ it is plausible that the longer the duration of dosing regimen, the more total AEs may be observed. However, due to the small number of RCTs for HCQ, it is not possible to determine the duration when HCQ would produce more AEs.

## Conclusions

Taken together, our data show that participants taking either CQ or HCQ experienced more mild and total AEs relative to placebo control. Precautionary measures should be taken when giving these medications for their therapeutic impact.

## Data Availability

Corresponding authors can provide data upon request.

## Acknowledgments

This work was supported by American Heart Association Predoctoral Award 18PRE34030199 (LR); Postdoctoral Fellowship from NIH F32 HL149288 (PNT); NIH R01 HL085727, HL085844, HL137228, and S10 RR033106, Research Award from the Rosenfeld Foundation, VA Merit Review Grant I01 BX000576 and I01 CX001490 (NC).

## Authors’ Contributions

LR and PNT designed the study. LR and PNT screened and evaluated studies. LR performed statistical analyses. SY checked studies included. PNT and SY checked statistical analyses. LR, WX, JO, and PNT performed comprehensive characterization of studies. SY and NC provided expertise. LR, PNT, and NC wrote the manuscript.

